# Soft-Tissue versus Hematologic Primary Malignant Cardiac Tumors: Demographics and First-Course Treatment Patterns in the SEER Registry

**DOI:** 10.64898/2026.08.25.26361262

**Authors:** Zubin Mathew, Riya Mehta, Samuel Kim, Johanna Jeyaraj, Talal Asif

## Abstract

**Background:** Primary malignant cardiac tumors (PMCTs) are rare and histologically heterogeneous.

**Objective:** To compare demographics, specific ICD-O-3 morphologies, first-course treatment patterns, annual registered case counts, and unadjusted overall survival between soft-tissue and hematologic PMCTs.

**Methods:** We identified 730 PMCT cases diagnosed from 2000 to 2021 in SEER 18 (ICD-O-3 topography C38.0). Histologic lineage was assigned from ICD-O-3 morphology. Comparative analyses included soft-tissue (n=458) and hematologic (n=212) tumors. First-course variables were primary-site surgery, chemotherapy (yes versus no/unknown), and radiotherapy (radiation versus none/unknown). Groups were compared with chi-square tests. Overall survival was estimated with Kaplan-Meier methods; follow-up was truncated at 120 months.

**Results:** Soft-tissue PMCTs occurred predominantly at ages 45–64 years (67.9%), whereas hematologic PMCTs occurred predominantly at age ≥65 years (63.2%; p<0.001). Men comprised 59.9% of hematologic and 49.3% of soft-tissue cases (p=0.014). The leading soft-tissue morphology was hemangiosarcoma/angiosarcoma (ICD-O-3 9120/3; 201/458, 43.9%); synovial sarcoma accounted for 20/458 cases (4.4%). Diffuse large B-cell lymphoma, NOS, accounted for 131/212 hematologic tumors (61.8%). Any primary-site surgery was recorded in 66.6% of soft-tissue versus 15.6% of hematologic cases (p<0.001). Chemotherapy was recorded in 67.5% versus 51.1% (p<0.001), and radiotherapy in 9.0% versus 20.5% (p<0.001). In exploratory Kaplan-Meier analyses, hematologic patients with recorded chemotherapy had higher unadjusted 120-month overall survival than those without recorded chemotherapy (42.0% versus 12.2%; log-rank p=7.5×10□□). Radiation-associated survival differences were not statistically significant in either lineage.

**Conclusions:** Soft-tissue and hematologic PMCTs have distinct age distributions, named histologies, and first-course treatment patterns in SEER. These findings describe registry coding and do not establish treatment effectiveness or population incidence.

**What is already known on this topic:** PMCTs are rare, biologically heterogeneous, and carry poor prognosis, with histologic lineage, rather than location or size, likely playing a critical role in prognostication. However, large population studies have rarely examined these tumors by specific histologic family, leaving demographic information, treatment, and prognosis across various subtypes poorly understood.

**What this study adds:** This manuscript is a secondary analysis of the same SEER 18, 2000-2021, ICD-O-3 site C38.0 cohort (n=730 after excluding 54 records without survival time) reported in our prior prognosis-group study. Our previous paper described overall demographics, literature-based 5-year prognosis strata, treatment frequencies, and Cox models in the pooled cohort. The present study does not recruit a new population. What is new is a lineage-based reorganization of the same cases: (1) separate demographic and first-course treatment profiles for soft-tissue versus hematologic tumors; (2) a complete ICD-O-3 morphology inventory, with angiosarcoma/hemangiosarcoma (9120/3) and DLBCL as the dominant named histologies; (3) annual registered case counts by lineage; and (4) exploratory lineage-stratified Kaplan-Meier overall survival comparisons by first-course chemotherapy and radiotherapy, with 95% confidence intervals and numbers at risk. Because SEER does not provide chemotherapy or radiotherapy start dates in this extract, these survival comparisons describe unadjusted associations and do not establish causal treatment effects.

**How this study might affect research, practice or policy:** By separating soft-tissue and hematologic PMCTs, this work supports histology-driven reporting and registry analyses rather than treating cardiac malignancies as a single entity. The lineage-specific treatment and survival patterns may inform hypothesis generation for prospective studies of cardiac sarcoma and cardiac lymphoma, but unadjusted SEER associations should not directly change clinical practice until treatment timing and confounding can be addressed.

## Introduction

Primary malignant cardiac tumors (PMCTs) are rare but necessary components of differential diagnoses for space-occupying lesions of the heart.^1^ The majority of primary cardiac tumors are benign and have shown an increasing incidence in recent years.^2^ Comprising only 5 to 6% of primary cardiac tumors, the malignant category has also seen an increased incidence over recent decades, possibly attributed to improved detection rather than true changes in prevalence.^2,3^ These cancerous tumors tend to have a poorer prognosis and heterogeneous presentation.^4^ To date, there have been no singular features that have been shown to be specific or sensitive in the diagnosis of PMCT.^3,5^

PMCTs reported prevalence ranges from 0.001% to 0.3%, and approximately 10% of all primary heart and pericardial tumors are malignant.^1^ Metastatic cardiac tumors are approximately 100 times more common than primary tumors.^6^ Younger patients tend to be affected more often by PMCTs, with the mean age at diagnosis around 44 years.^7^ Both males and females are affected equally, and racial differences in survival outcomes have not been observed in the literature.^7^ PMCT incidence has increased from approximately 25 cases per year between the late 20th century and the early 2000s to around 47 cases per year in the more recent decade.^5^ PMCTs present with vague cardiopulmonary symptoms causing late presentation; the most frequent presenting symptom is dyspnea.^7^

The most common PMCTs include sarcomas, lymphomas, and a diverse range of other histopathologies.^8^ PMCTs comprise a biologically heterogeneous group of neoplasms in which histologic lineage appears to be an important determinant of prognosis.^9^ Because these tumors vary wildly in growth, metastasis, and treatment response, they cannot be treated as a single category. Instead, the specific histologic lineage, rather than the cardiac location itself, is the most critical factor determining a patient’s prognosis.^8^

From a clinical standpoint, histopathology strongly determines management. Sarcoma-predominant tumors are more often approached with surgical resection when feasible, while hematologic malignancies may depend more heavily on systemic therapy for broader disease control.^8^ These distinctions are important because resectability and treatment modality do not affect all PMCTs in the same way. A histology-centered approach would offer a more precise framework for interpreting prognosis and may help clarify why some patients benefit from aggressive local therapy while others have a course driven primarily by systemic spread.

Population-based studies have often pooled cardiac malignancies or grouped them by literature-derived prognosis rather than by lineage. We previously described SEER PMCT cases diagnosed from 2000 to 2021 using literature-based 5-year prognosis strata.^10^ The present analysis uses that same cohort to compare soft-tissue and hematologic tumors as distinct histologic families. The aims were to (1) inventory ICD-O-3 morphologies within each lineage, (2) compare demographic and first-course treatment patterns, (3) display annual registered case counts, and (4) describe exploratory overall survival by first-course chemotherapy and radiotherapy within each lineage using Kaplan-Meier methods. Treatment comparisons are presented as unadjusted registry associations and are not interpreted as evidence of therapeutic benefit.

## Methods

### Study Population and Cohort Selection

We queried SEER*Stat (November 2023 submission) for SEER 18 cases diagnosed from 2000 to 2021 with primary site ICD-O-3 C38.0 (heart).^11^ After exclusion of 54 records missing survival time, 730 patients remained. Demographic variables were sex; race/ethnicity recoded as White, Black, or Other; rural-urban continuum (urban versus rural); and age at diagnosis categorized as youth (<45 years), middle (45-64 years), or elderly (≥65 years). Neighborhood median household income, inflation-adjusted to 2022, was dichotomized at the 2022 US median ($75,000). Clinical variables included ICD-O-3 histology, tumor size summary (available 2016 onward), and SEER combined metastasis indicators at diagnosis for bone, brain, liver, and lung (available 2010 onward).

Each ICD-O-3 morphology code was assigned to one of four lineages, soft tissue, hematologic, embryonal, or other, by matching the code to standard ICD-O-3 histologic families (mesenchymal/sarcoma, lymphoid/hematopoietic, embryonal/germ-cel-related, or epithelial/mesothelial/miscellaneous). The complete code list is provided in Supplementary Table 1. Comparative analyses were restricted to soft-tissue (n=458) and hematologic (n=212) tumors. Embryonal (n=7) and other (n=53) tumors are described only in the histology inventory because of small size and heterogeneity.

### Treatment Variables

Treatment covariates reflect first-course therapy only. Subsequent therapy after progression or recurrence is not captured. Surgery of the primary site was recoded as none (blank SEER surgery code), any documented primary-site procedure, or unknown/death-certificate only. Detailed SEER surgery codes are listed in Supplementary Table 2. Chemotherapy is the SEER recode “yes” versus “no/unknown.” Radiotherapy is “radiation” versus “no/unknown.” SEER cannot reliably separate confirmed non-receipt from unknown receipt, so patients without a “yes” marker are not described as untreated. Sequence and intent are not available, so chemotherapy is reported as recorded receipt rather than as adjuvant or neoadjuvant therapy. A four-level first-course variable was created: neither chemotherapy nor radiotherapy documented; chemotherapy only; radiotherapy only; and both.

### Outcomes and statistical analysis

The primary outcome was overall survival from diagnosis to death from any cause or last known follow-up. Vital status was derived from the SEER cause-of-death recode: patients coded as alive were censored; all other cause-of-death codes were treated as events. Follow-up time was truncated at 120 months, with patients alive beyond 120 months administratively censored at 120 months. Kaplan-Meier curves were constructed separately within each lineage for first-course chemotherapy (yes versus no/unknown) and radiotherapy (radiation versus none/unknown). Curves display 95% confidence intervals (Greenwood method), censoring marks, and numbers at risk at 0, 20, 40, 60, 80, 100, and 120 months. Group differences were tested with the log-rank test. Median overall survival and survival probabilities at 30, 60, and 120 months are reported with 95% confidence intervals in the Results. One hematologic chemotherapy Kaplan-Meier figure is shown (Figure 3); remaining comparisons are summarized numerically because of figure limits and small treated subgroups. These observational comparisons are vulnerable to confounding by indication, immortal-time bias, and guarantee-time bias because treatment start dates are not available in SEER; they do not establish that chemotherapy or radiotherapy improves survival.

Categorical variables are presented as counts and percentages. Differences between soft-tissue and hematologic cohorts were compared with chi-square tests; unknown surgery (n=4) was excluded from the surgery comparison. Two-sided p<0.05 was considered statistically significant. Analyses were exploratory and were not adjusted for multiple comparisons; we emphasize absolute percentages and numbers at risk alongside p-values. Annual counts from 2000 to 2021 are registered cases, not age-standardized incidence rates, because SEER population denominators were not available. Missingness is summarized in Supplementary Table 3. This observational registry report follows STROBE guidance.

### Ethics

This study used de-identified data from the publicly available SEER Program. The dataset contains no identifiable patient information and involves no contact with participants. The study was considered exempt from institutional review board review, and the requirement for informed consent was waived, consistent with the Declaration of Helsinki.

## Results

### Demographics

Table 1 shows demographic and treatment characteristics. Age distribution differed by lineage (p<0.001). Hematologic tumors were concentrated in patients aged ≥65 years (134/212, 63.2%), with only six patients younger than 45 years (2.8%). Soft-tissue tumors were concentrated at ages 45-64 years (311/458, 67.9%); 48 patients (10.5%) were younger than 45 years and 99 (21.6%) were aged ≥65 years. Hematologic cases were more often male (59.9% versus 49.3%; p=0.014). Race/ethnicity recodes did not differ significantly (p=0.21). Most patients in both lineages lived in urban counties and in counties with above-median household income; these distributions did not differ between lineages (p=0.18 and p=0.44).

**Table 1.** Demographics and first-course treatment for hematologic versus soft-tissue primary malignant cardiac tumors, SEER 18, 2000-2021.

| Variable | Hematologic (n=212) | Soft tissue (n=458) | p |
| --- | --- | --- | --- |
| Age group |  |  | <0.001 |
| $\geq 65$ years | 134 (63.2) | 99 (21.6) | |
| 45-64 years | 72 (34.0) | 311 (67.9) |  |
| <45 years | 6 (2.8) | 48 (10.5) |  |
| Sex |  |  | 0.014 |
| Male | 127 (59.9) | 226 (49.3) |  |
| Female | 85 (40.1) | 232 (50.7) |  |
| Race |  |  | 0.21 |
| White | 134 (63.2) | 264 (57.6) |  |
| Other | 63 (29.7) | 144 (31.4) |  |
| Black | 15 (7.1) | 50 (10.9) |  |
| County median household income |  |  | 0.44 |
| $\geq \$75,000$ | 160 (75.5) | 331 (72.3) | |
| <\$75,000 | 52 (24.5) | 127 (27.7) | |
| Rural-urban continuum |  |  | 0.18 |
| Urban | 197 (92.9) | 409 (89.3) |  |
| Rural | 15 (7.1) | 49 (10.7) |  |
| Primary-site surgery |  |  | <0.001* |
| Any documented surgery | 33 (15.6) | 305 (66.6) |  |
| None | 177 (83.5) | 151 (33.0) |  |
| Unknown/death certificate | 2 (0.9) | 2 (0.4) |  |
| Chemotherapy recode |  |  | <0.001 |
| Yes | 143 (67.5) | 234 (51.1) |  |
| No/unknown | 69 (32.5) | 224 (48.9) |  |
| Radiotherapy recode |  |  | <0.001 |
| Yes | 19 (9.0) | 94 (20.5) |  |
| No/unknown | 193 (91.0) | 364 (79.5) |  |
| Chemotherapy and radiotherapy |  |  | <0.001 |
| Neither documented | 67 (31.6) | 197 (43.0) |  |
| Chemotherapy only | 126 (59.4) | 167 (36.5) |  |
| Radiotherapy only | 2 (0.9) | 27 (5.9) |  |
| Both | 17 (8.0) | 67 (14.6) |  |

*Values are n (%). P values are chi-square tests comparing lineages.*
*\*Surgery comparison excludes unknown/death certificate (n=4). Income is county-level, dichotomized at* *the 2022 US median, not individual socioeconomic status. Chemotherapy and radiotherapy use SEER yes* *versus no/unknown recodes. Detailed surgery codes are in Supplementary Table 2.*

### Histologic inventory

Table 2 and Supplementary Table 1 summarize ICD-O-3 morphologies. Among 458 soft-tissue PMCTs, the most common morphology was hemangiosarcoma (ICD-O-3 9120/3), the ICD-O-3 term corresponding to angiosarcoma, in 201 patients (43.9%). This was followed by sarcoma, not otherwise specified (NOS) (8800/3; 8.7%), giant cell sarcoma (8802/3; 8.1%), spindle cell sarcoma (8801/3; 5.0%), and leiomyosarcoma, NOS (8890/3; 4.8%). Synovial sarcoma (9040/3, 9041/3, and 9043/3 combined) accounted for 20 cases (4.4%).

**Table 2.** Leading ICD-O-3 morphologies by histologic lineage.

| Lineage | ICD-O-3 morphology | n | % of lineage |
| --- | --- | --- | --- |
| Soft tissue (n=458) | 9120/3 Hemangiosarcoma (angiosarcoma) | 201 | 43.9 |
|  | 8800/3 Sarcoma, NOS | 40 | 8.7 |
|  | 8802/3 Giant cell sarcoma | 37 | 8.1 |
|  | 8801/3 Spindle cell sarcoma | 23 | 5.0 |
|  | 8890/3 Leiomyosarcoma, NOS | 22 | 4.8 |
|  | 9040/3, 9041/3, 9043/3 Synovial sarcoma (all) | 20 | 4.4 |
| Hematologic (n=212) | 9680/3 DLBCL, NOS | 131 | 61.8 |
|  | 9591/3 Non-Hodgkin lymphoma, NOS | 25 | 11.8 |
|  | 9678/3 Primary effusion lymphoma | 20 | 9.4 |
|  | 9590/3 Malignant lymphoma, NOS | 7 | 3.3 |
| Embryonal (n=7) | 9080/3 Teratoma, malignant, NOS | 4 | 57.1 |
|  | 9473/3 Primitive neuroectodermal tumor | 2 | 28.6 |
|  | 8963/3 Malignant rhabdoid tumor | 1 | 14.3 |
| Other (n=53) | 9052/3 Epithelioid mesothelioma, malignant | 15 | 28.3 |
|  | 8140/3 Adenocarcinoma, NOS | 9 | 17.0 |
|  | 9050/3 Mesothelioma, malignant | 5 | 9.4 |

The complete ICD-O-3 inventory is provided in Supplementary Table 1.

Among 212 hematologic PMCTs, diffuse large B-cell lymphoma, NOS (9680/3) predominated (131/212, 61.8%), followed by non-Hodgkin lymphoma, NOS (9591/3; 11.8%) and primary effusion lymphoma (9678/3; 9.4%). Embryonal tumors (n=7) comprised malignant teratoma, NOS (n=4), primitive neuroectodermal tumor (n=2), and malignant rhabdoid tumor (n=1). In the other category (n=53), epithelioid mesothelioma (9052/3; 28.3%), adenocarcinoma, NOS (8140/3; 17.0%), and malignant mesothelioma (9050/3; 9.4%) were most frequent.

### Registered Case Counts Over Time

Figure 1 shows annual registered counts for the full cohort (n=730) from 2000 through 2021. Year-to-year counts fluctuated in both lineages. Period totals were 185 cases in 2000-2006, 235 in 2007-2013, and 310 in 2014-2021. Hematologic counts were 53, 69, and 90 across those periods; soft-tissue counts were 115, 150, and 193. Hematologic tumors comprised 28.6% of registered PMCTs in 2000-2006 and 29.0% in 2014-2021. Absolute counts were higher in later years in both lineages; the hematologic share of the cohort was stable. These figures are case counts, not incidence rates.

**Figure 1.**
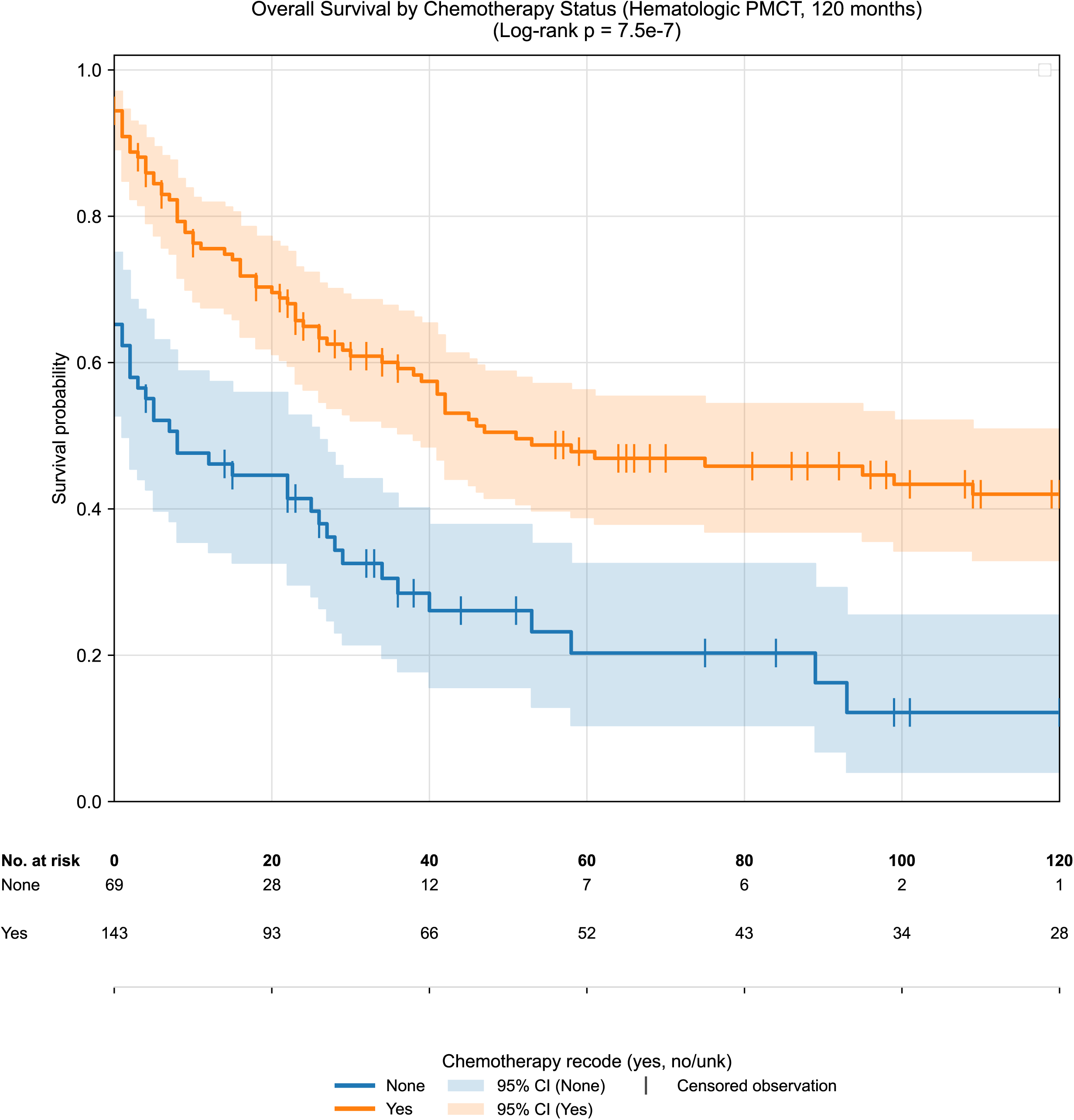
Annual number of registered primary malignant cardiac tumors in SEER 18, 2000-2021, by histologic lineage. Soft-tissue (n=458) and hematologic (n=212) tumors are shown separately; embryonal and other tumors are combined (n=60). Counts are registered cases and are not population-based incidence rates. Source: SEER 18, ICD-O-3 site C38.0.

### First-Course Treatment

Figure 2 displays the first-course treatment mix. Any documented primary-site surgery was recorded in 305/458 soft-tissue tumors (66.6%) versus 33/212 hematologic tumors (15.6%; p<0.001). No surgery was recorded in 33.0% and 83.5%, respectively; surgery was unknown on the death certificate in two patients in each group. When excisional biopsy was excluded, a resection-like procedure remained recorded in 222/458 soft-tissue cases (48.5%) and 23/212 hematologic cases (10.8%).

**Figure 2.**
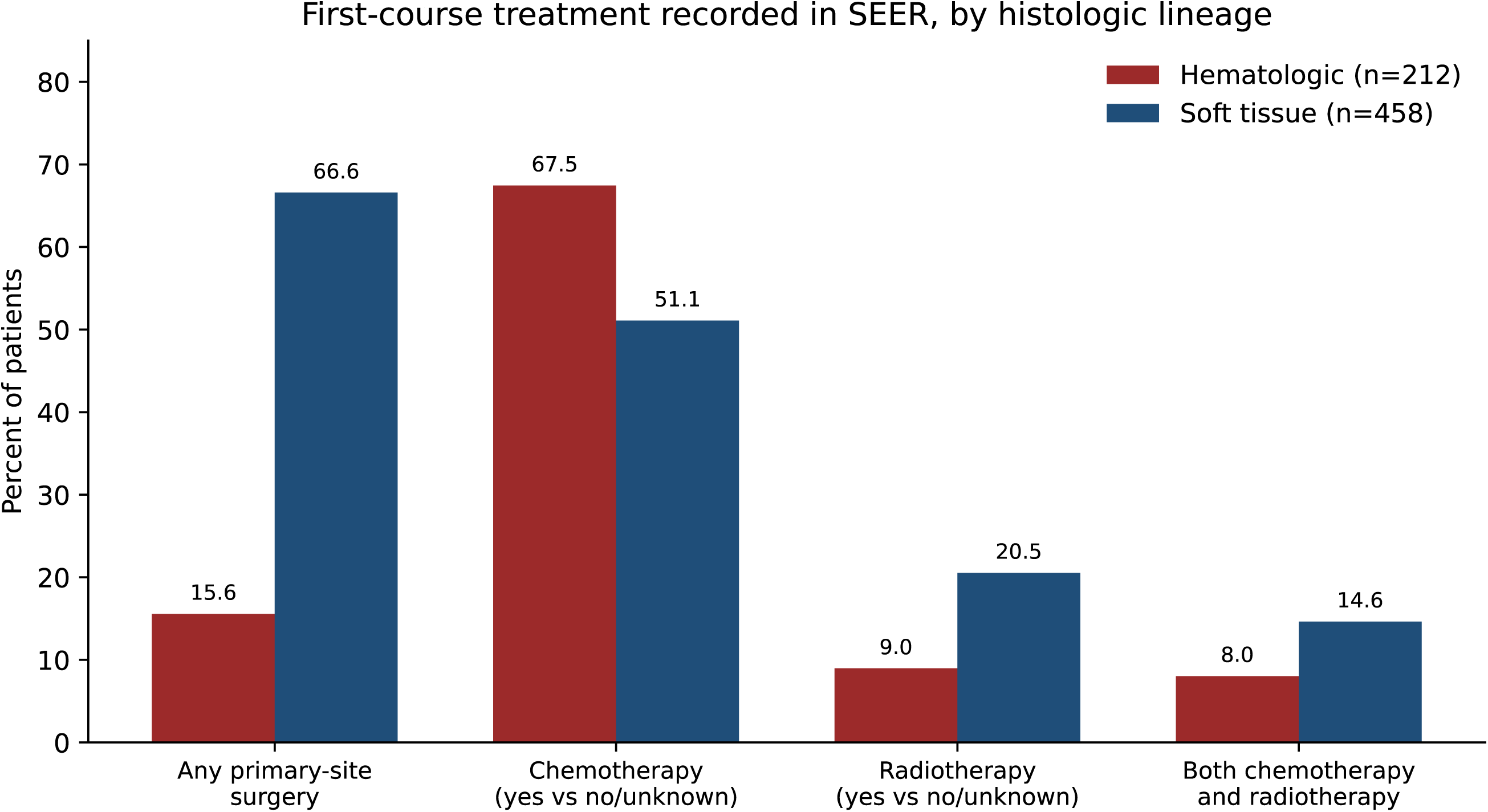
First-course treatment recorded in SEER among hematologic (n=212) and soft-tissue (n=458) primary malignant cardiac tumors. Bars show the percent of patients with any documented primary-site surgery, chemotherapy recorded as yes (versus no/unknown), radiotherapy recorded as yes (versus no/unknown), and both chemotherapy and radiotherapy recorded. Percentages describe registry coding and are not treatment-effect estimates.

**Figure 3:**
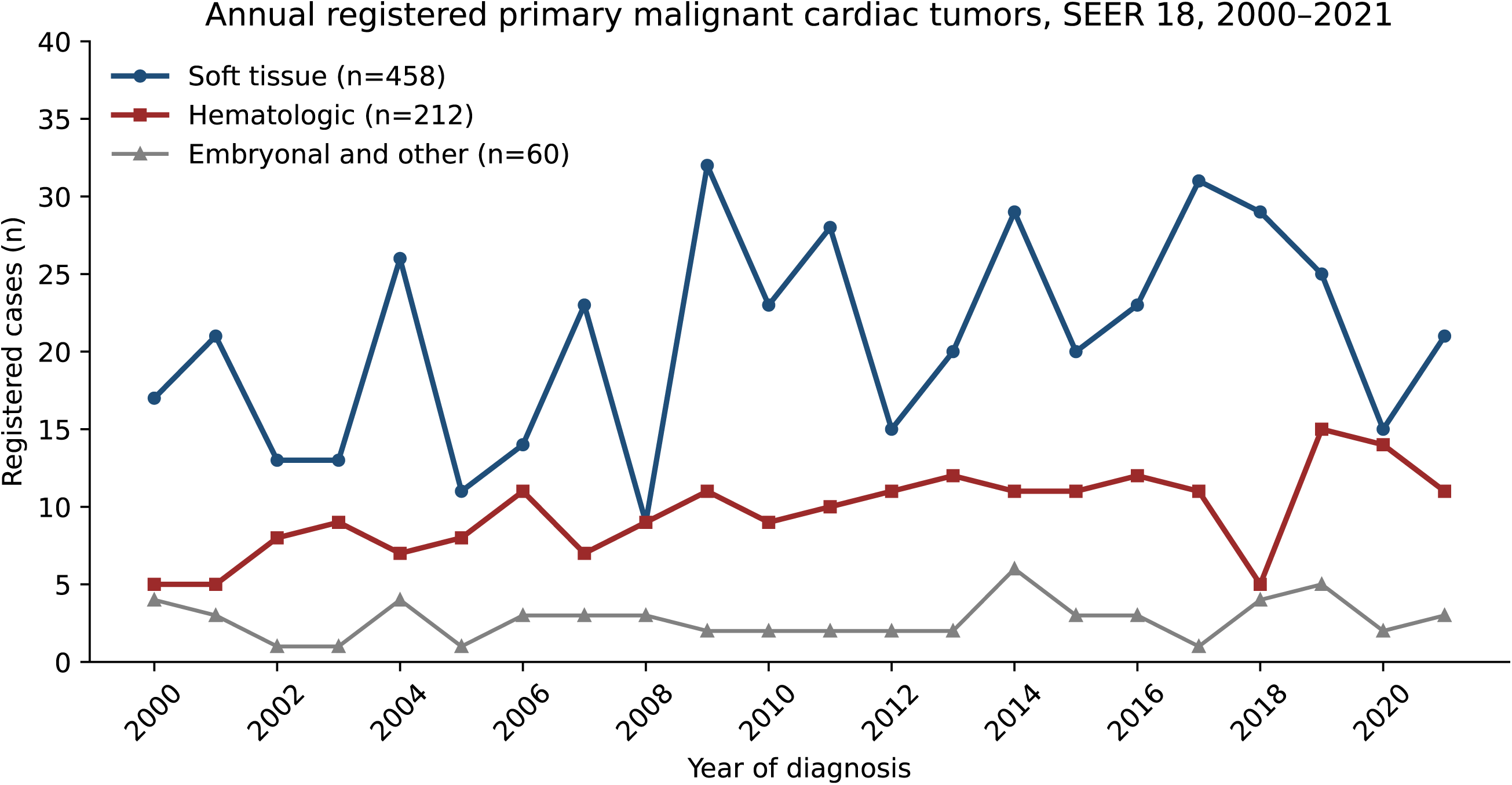
Unadjusted Kaplan-Meier Overall Survival estimates by First-course Chemotherapy Status in Hematologic Primary Malignant Cardiac Tumors. Figure 3. Kaplan-Meier overall survival by first-course chemotherapy status among hematologic primary malignant cardiac tumors (n=212), truncated at 120 months. Shaded bands denote group-specific 95% confidence intervals; vertical tick marks indicate censored observations. Numbers at risk are shown below the x-axis at 20-month intervals. Log-rank p=7.5×10^-7^. This observational comparison does not establish treatment benefit.

Chemotherapy (yes versus no/unknown) was recorded in 143/212 hematologic tumors (67.5%) and 234/458 soft-tissue tumors (51.1%; p<0.001). Radiotherapy was recorded in 19/212 hematologic tumors (9.0%) and 94/458 soft-tissue tumors (20.5%; p<0.001). Combined first-course chemotherapy and radiotherapy was recorded in 17 hematologic patients (8.0%) and 67 soft-tissue patients (14.6%). In the hematologic cohort, 17 of the 19 patients with radiotherapy also had chemotherapy recorded; only two had radiotherapy without chemotherapy. In soft tissue tumors, neither modality was documented in 197 patients (43.0%), chemotherapy only in 167 (36.5%), radiotherapy only in 27 (5.9%), and both in 67 (14.6%). These percentages describe SEER first-course flags and do not establish therapeutic benefit.

### Overall survival by first-course treatment

Exploratory Kaplan-Meier analyses compared overall survival by first-course chemotherapy and radiotherapy within each lineage, with follow-up truncated at 120 months. One hematologic chemotherapy Kaplan-Meier figure is shown as Figure 3. Hematologic tumors (n=212): Among patients with chemotherapy recorded as yes (n=143) versus no/unknown (n=69), median overall survival was 51 months (95% CI 39-63) versus 8 months (95% CI 4-12; log-rank p=7.5×10^-7^). Survival probabilities at 30, 60, and 120 months were 60.9% (95% CI 52.1-68.6%), 47.8% (95% CI 37.9-55.4%), and 42.0% (95% CI 32.9-50.9%) in the chemotherapy-yes group versus 32.5% (95% CI 21.5-44.1%), 20.3% (95% CI 10.4-32.5%), and 12.2% (95% CI 3.9-25.5%) in the no/unknown group. Numbers at risk at 120 months were 28 and 1, respectively (Figure 3). A substantial proportion of patients without recorded chemotherapy died at or immediately after diagnosis (24/69 deaths at month 0), indicating that many untreated patients did not survive long enough for therapy to be initiated.

Among hematologic patients, radiotherapy was recorded in only 19 patients (beam radiation or recommended, unknown if administered) versus 191 with none/unknown. Median overall survival was 75 months (95% CI 41-97) with radiotherapy versus 29 months (95% CI 23-37) without (log-rank p=0.23).

Ten-year survival estimates were 41.9% (95% CI 20.9-61.7%) versus 32.4% (95% CI 24.7-40.3%), but only eight irradiated patients remained at risk at 120 months; these estimates are therefore imprecise and not statistically significant.

Soft-tissue tumors (n=458): Chemotherapy recorded as yes (n=234) versus no/unknown (n=223) was associated with longer median overall survival (16 versus 3 months; log-rank p=3.6×10^-7^) and higher 30-month survival (21.9% [95% CI 16.5-27.7%] versus 16.9% [95% CI 12.2-22.4%]), but 120-month survival converged (4.6% [95% CI 1.6-10.1%] versus 5.4% [95% CI 2.7-9.4%]) with only three and seven patients at risk, respectively. Radiotherapy was recorded in 99 soft-tissue patients versus 358 with none/unknown (median overall survival 15 versus 10 months; log-rank p=0.20). At 120 months, only one irradiated patient remained at risk. These unadjusted comparisons do not establish that chemotherapy or radiotherapy improves survival in either lineage.

### Missing data

Age, sex, race recode, year, income, rural-urban status, ICD-O-3 morphology, and lineage were complete. Tumor size summary (2016+) was missing in 500/730 (68.5%), including 144/212 hematologic and 314/458 soft-tissue cases, and was not analyzed. Combined metastasis variables (2010+) were missing in approximately half of the cohort and in 74% of hematologic cases; they were not used for lineage comparisons. Blank chemotherapy and radiotherapy fields are no/unknown recodes rather than ordinary item nonresponse (Supplementary Table 3).

## Discussion

In this SEER cohort of 730 PMCTs, soft-tissue and hematologic tumors had different age and sex profiles, different leading morphologies, and different first-course treatment patterns. Hematologic PMCTs clustered in older men. Soft-tissue PMCTs clustered in middle-aged adults of both sexes. These demographic differences were larger for age (Table 1) and more modest for sex. Race recode, county income, and rural-urban residence did not differ by lineage and should not be read as evidence of differential access or underdiagnosis, because the analysis has no population denominators and income is an area-level measure.

The Histology Inventory in Table 2 is consistent with the clinical literature on cardiac sarcoma and cardiac lymphoma.^7,9^ Hemangiosarcoma/angiosarcoma accounted for 43.9% of soft-tissue tumors, whereas synovial sarcoma accounted for 4.4%. Other SEER cardiac-sarcoma series likewise find angiosarcoma to be the leading subtype and synovial sarcoma to be uncommon.^12–14^ Named hematologic disease was dominated by DLBCL (61.8%), with a smaller primary effusion lymphoma subset (9.4%), a distribution that is biologically plausible for cardiac and pericardial lymphoma.^13^ Discussion of histology-guided care for these tumors is therefore better linked to cardiac sarcoma and cardiac lymphoma series than to unrelated solid tumors.^8,12,13^

Figure 2 shows the treatment dichotomy that follows from that histology. Surgery was the modality that most clearly separated the two lineages, recorded in two-thirds of soft-tissue tumors and in a small minority of hematologic tumors. Chemotherapy was the predominant coded modality among hematologic cases (67.5%) and was still recorded in about half of soft-tissue cases. Radiotherapy was uncommon in both groups and particularly uncommon in hematologic tumors (9.0%), where 17 of 19 irradiated patients also had chemotherapy recorded. Combined chemotherapy and radiotherapy was therefore essentially a subset of the hematologic radiotherapy group (n=17) and a modest subset of soft-tissue cases (n=67). These patterns align with usual approaches to cardiac lymphoma and cardiac sarcoma.^8,12,13^ They do not identify treatment sequence or intent, they mix unknown receipt with non-receipt in the no/unknown category, and they cannot be interpreted as evidence that surgery, chemotherapy, or radiotherapy improves survival.

Figure 3 illustrates the hematologic chemotherapy comparison requested for revision, including 95% confidence intervals, censoring marks, and numbers at risk. The apparent survival advantage among patients with recorded chemotherapy likely reflects both confounding by indication and the high burden of early mortality in the no/unknown group. Similarly, the small hematologic radiotherapy subgroup (n=19) and sparse long-term numbers at risk preclude strong inference about durable radiotherapy benefit. In soft-tissue tumors, chemotherapy and radiotherapy comparisons showed similar limitations: statistically detectable early differences for chemotherapy but overlapping long-term estimates with very few patients remaining at risk. Accordingly, we describe treatment-associated survival patterns as exploratory registry associations rather than evidence that systemic therapy improves outcome.

Each broad category, of either hematologic or soft tissue comprises a wide array of specific histopathologic subtypes. This diversity is clinically important because different histologic subtypes carry distinct biologic behaviors, prognoses, treatment implications, and management of PMCTs and thus treatment is increasingly guided by histology rather than by a single unified approach to cardiac malignancy.^14,15^ Given the extreme rarity of these tumors and the absence of prospective randomized trials, treatment selection must often be guided by histologic lineage rather than by a uniform framework for cardiac malignancy.^8^ This subtype-specific approach is clinically meaningful because existing series suggest that prognosis and treatment responsiveness vary considerably across histologies, with lymphoma demonstrating the clearest potential for durable response to anthracycline-based immunochemotherapy and cardiac sarcomas generally being managed according to principles derived from their soft tissue counterparts.^16^ In this context, histopathologic classification is not merely descriptive but may directly influence whether patients are directed toward surgery, systemic therapy, or multimodality treatment, thereby affecting the likelihood of timely and appropriate intervention.

Figure 1 places these cases in a timeline. Registered counts were higher in 2014-2021 than in 2000-2006 for both lineages, with substantial year-to-year variation. The hematologic fraction of all PMCTs remained near 29% throughout. Because Figure 1 does not incorporate SEER population denominators, the upward drift in counts is not a demonstration of rising incidence. The dataset also contains no etiologic exposures (HIV status, immunosuppression, occupation, or germline data), so the figure cannot be used to infer risk factors. Improved cardiac imaging is a possible contributor to ascertainment, but that hypothesis is not tested here.

This analysis uses the same SEER 18, 2000-2021, C38.0 extract as our previous prognosis-group study (n=730 after exclusion of 54 records without survival time).10 That report described the pooled cohort and literature-based survival strata. The present report reorganizes those cases by histologic lineage and adds the named-morphology inventory, the lineage-specific treatment table, the chemotherapy/radiotherapy cross-classification, annual count figures, and exploratory lineage-stratified overall survival comparisons with 95% confidence intervals and numbers at risk.

### Implications and Future Directions

The lineage-specific patterns in this SEER cohort highlight that PMCTs should not be analyzed or managed as a uniform entity. Soft-tissue and hematologic PMCTs exhibit distinct demographic clustering, predominant named histologies (angiosarcoma versus DLBCL), and initial management approaches, supporting routine lineage-stratified reporting in future registry work. First-course treatment patterns reflect expected clinical practice, such as surgical predominance in soft-tissue tumors and chemotherapy in hematologic cases, but represent registry documentation rather than validated therapeutic efficacy. Unadjusted Kaplan-Meier comparisons must be interpreted cautiously: patients who receive therapy must survive long enough and remain clinically suitable for treatment, whereas the no/unknown groups include patients who may have died before therapy could be delivered. Future studies require registry extracts with exact treatment start dates to support time-dependent Cox models, landmark analyses, or other designs that address treatment timing. Prospective multi-institutional registries and standardized pathology review remain essential to evaluate sequencing, confounding, and rare-subgroup stability.

## Limitations

This study has several limitations inherent to the SEER database and the available analytic extract. First, although vital status could be inferred from cause-of-death coding, treatment start dates were unavailable; Kaplan-Meier comparisons of treated versus untreated patients therefore remain vulnerable to immortal-time bias, guarantee-time bias, and unmeasured confounding by indication. Observed separation of survival curves does not prove treatment benefit and may partly reflect early mortality in patients who never received therapy. Second, radiotherapy subgroup analyses were underpowered, only 19 hematologic patients received beam radiotherapy, and long-term estimates with very small numbers at risk are unstable and should not be interpreted as durable treatment effects. Third, SEER captures first-course therapy only, records chemotherapy and radiotherapy as “yes” or “radiation” versus “no/unknown,” and lacks data on regimen details, radiation fields, surgical margin status, and subsequent therapies at disease progression. Fourth, temporal analyses describe annual registered case counts rather than age-standardized population incidence rates, and geographic/income distributions reflect county-level ecological data rather than individual-level socioeconomic status. Fifth, tumor size (missing in 68.5%) and metastasis indicators at diagnosis (missing in ∼50%) were too incomplete for primary comparative adjustment. Finally, lineage assignments relied on ICD-O-3 coding without central pathology review, and findings reflect SEER registry participation, which may not fully generalize to all healthcare settings or uncaptured cases presenting as sudden cardiac death.

## Conclusions

In SEER, soft-tissue PMCTs are predominantly middle-aged patients. The leading named morphology is angiosarcoma/hemangiosarcoma, and primary-site surgery is the first-course modality most often recorded. Hematologic PMCTs are predominantly older men. The leading named morphology is DLBCL, and chemotherapy is the first-course modality most often recorded. Annual registered counts increased in both lineages while the hematologic share of cases remained stable. Exploratory Kaplan-Meier analyses showed higher unadjusted overall survival among hematologic patients with recorded chemotherapy, but treatment comparisons are confounded and do not establish therapeutic benefit. These findings describe who is recorded in the registry and which first-course modalities are coded. They do not define optimal therapy or population incidence.

## List of Abbreviations

PMCT: Primary Malignant Cardiac Tumor
SEER: Surveillance, Epidemiology, and End Results
ICD: International Classification of Disease
OS: Overall Survival
KM: Kaplan Meier
DLBCL: Diffuse Large B-cell Lymphoma
NOS: Not Otherwise Specified
CLL: chronic lymphocytic leukemia
SLL: small lymphocytic lymphoma
PNET: primitive neuroectodermal tumor

## Declarations

### Ethics approval and consent to participate

This study utilized de-identified data from the publicly available Surveillance, Epidemiology, and End Results (SEER) Program database. As the dataset contains no identifiable patient information and involves no direct interaction with human participants, this study was considered exempt from Institutional Review Board review and the requirement for informed consent was waived, consistent with the ethical principles outlined in the Declaration of Helsinki.

### Consent for publication

Informed consent was waived because this study used publicly available, de-identified data from the Surveillance, Epidemiology, and End Results (SEER) Program database, which contains anonymized cancer registry information collected for public health surveillance. No identifiable patient information was accessed.

### Availability of data and materials

The data analyzed in this study was obtained from the SEER database. Requests for access to these datasets should be directed to https://seer.cancer.gov/data/access.html

### Author Contributions

Z.M.^*✝^: Conceptualization, Investigation, Writing-original draft, Writing-review, editing, project administration, supervision.

R.M.^*✝^: Investigation, data curation and formal analysis, Writing-original draft, Writing-review, editing.

Both authors, Z.M. and R.M., contributed equally to this work and should be considered as co-first authors, if acceptable by the journal.

S.K.: Data curation, formal analysis, resources, validation, writing original draft, review.

J.J.: Writing-original draft, Writing-review & editing.

T.A.: Supervision & project administration.

All authors read and approved the submitted version.

### Data Availability Statement

Data is publicly available and submitted with the manuscript as Supplemental Material.

### Guarantor Statement

Riya Mehta accepts full responsibility for the work and/or the conduct of the study, had access to the data, and controlled the decision to publish.

## References

1. Ghosh AK, Walker JM. Cardio-oncology. Br J Hosp Med. 2017;78(1):C11–C13. doi:10.12968/hmed.2017.78.1.C11

2. Cresti A, Chiavarelli M, Glauber M, et al. Incidence rate of primary cardiac tumors: a 14-year population study. J Cardiovasc Med. 2016;17(1):37. doi:10.2459/JCM.0000000000000059

3. MR Imaging of Cardiac Tumors | RadioGraphics. Accessed September 17, 2025. https://pubs.rsna.org/doi/10.1148/rg.255045721?url_ver=Z39.88-2003&rfr_id=ori:rid:crossref.org&rfr_dat=cr_pub%20%200pubmed

4. Leja MJ, Shah DJ, Reardon MJ. Primary Cardiac Tumors. Tex Heart Inst J. 2011;38(3):261.

5. Burke AP, Rosado-de-Christenson M, Templeton PA, Virmani R. Cardiac fibroma: clinicopathologic correlates and surgical treatment. J Thorac Cardiovasc Surg. 1994;108(5):862–870.

6. Mc Allister HA. Primary tumors and cysts of the heart and pericardium. Curr Probl Cardiol. 1979;4(2):1–51. doi:10.1016/0146-2806(79)90008-2

7. Oliveira GH, Al-Kindi SG, Hoimes C, Park SJ. Characteristics and Survival of Malignant Cardiac Tumors. Circulation. 2015;132(25):2395–2402. doi:10.1161/CIRCULATIONAHA.115.016418

8. Shiba M, Nishiyama M, Sotomi Y, et al. Long-Term Outcomes of Primary Cardiac Tumors by Histological Subtype. Circ Rep. 8(3):472–478. doi:10.1253/circrep.CR-25-0234

9. Isobe S, Murohara T. Editorial: Cardiac tumors: Histopathological aspects and assessments with cardiac noninvasive imaging. J Cardiol Cases. 2015;12(2):37–38. doi:10.1016/j.jccase.2015.04.008

10. Mehta R, Mathew Z, Kim S, Jeyaraj J, Asif T. Prognostic trends of malignant cardiac tumors: insights from the Surveillance, Epidemiology, and End Results (SEER) registry. Explor Cardiol. 2026;4:1012107. doi:10.37349/ec.2026.1012107

11. SEER Incidence Data - SEER Data & Software. SEER. Accessed August 23, 2025. https://seer.cancer.gov/data/index.html

12. Kassi M, Polsani V, Schutt RC, et al. Differentiating benign from malignant cardiac tumors with cardiac magnetic resonance imaging. J Thorac Cardiovasc Surg. 2019;157(5):1912–1922.e2. doi:10.1016/j.jtcvs.2018.09.057

13. Hofmann WK, Trumpp A, Müller-Tidow C. Therapy resistance mechanisms in hematological malignancies. Int J Cancer. 2023;152(3):340–347. doi:10.1002/ijc.34243

14. Ravi V, Reardon M. Commentary: Primary cardiac sarcoma—Systemic disease requires systemic therapy. J Thorac Cardiovasc Surg. 2019; 161, 120–121. doi:10.1016/j.jtcvs.2019.10.087

15. Hasan S, Witten J, Collier P, et al. Outcomes after resection of primary cardiac sarcoma J Thorac Cardiovasc Surg, 2021; 8, 384–390. doi:10.1016/j.xjon.2021.08.038

16. Zhang PJ, Brooks JS, Goldblum JR, et al. Primary cardiac sarcomas: a clinicopathologic analysis of a series with follow-up information in 17 patients and emphasis on long-term survival. Hum Pathol. 2008;39(9):1385–1395. doi:10.1016/j.humpath.2008.01.019

